# Comparing Gleason Pattern 4 Measurement Approaches on Prostate Biopsy Using Machine Learning: A Proof-of-Principle Study

**DOI:** 10.64898/2026.04.23.26351615

**Authors:** Matei M. Buzoianu, Rebecca Yu, Melissa Assel, Alican Bozkurt, Hamed Aghdam, Samson W. Fine, Andrew J. Vickers

## Abstract

**Objective:** To demonstrate the proof of principle that machine learning (ML) can be used to quantify Gleason Pattern (GP) 4 on digitized biopsy slides using multiple measurement approaches, allowing direct comparison of their prognostic performance.

**Methods:** We assembled a convenience sample of 726 patients with grade group 2-4 prostate cancer on systematic biopsy who underwent radical prostatectomy between 2014 and 2023. Digitized biopsy slides were analyzed using a machine-learning algorithm (PAIGE-AI) to quantify GP4 using multiple measurement approaches, particularly with respect to how gaps between cancer foci (“interfocal stroma”) were handled. GP4 extent was quantified using linear measurements or a pixel-based area metric. Discrimination of each GP4 quantification approach, along with Grade Group (GG), was assessed for adverse radical prostatectomy pathology and biochemical recurrence.

**Results:** We identified 15 different quantification approaches and observed differences between their discrimination. The highest discrimination was in the pixel-counting method (AUC 0.648). GP4 quantification outperformed GG for predicting adverse pathology (AUC 0.627 vs 0.608). Amount of GP3 was non-predictive once GP4 was known. These findings were consistent for BCR.

**Conclusions:** We were able to measure slides using 15 distinct measurement approaches and replicated prior findings using ML to quantify GP4. Our findings support the use of ML as a research tool to compare different GP4 quantification approaches. We intend to use our method on larger cohorts to determine with which measurement approach best predicts oncologic outcome.

## Introduction

The Gleason Score is the primary determinant of management decisions in localized prostate cancer, normally expressed as the Grade Group (GG). GG1 is indolent [1] and is typically managed by active surveillance; GG5 has the poorest prognosis and is treated aggressively. The largest group, and the one in which treatment decisions are most challenging, is GG2–4. Risk stratification for such men is based on the ratio between Gleason pattern (GP) 4 and GP3. Use of ratios in unusual in oncology and leads to some counterintuitive findings [2]. For instance, a patient with a substantial burden of GP4, and a slightly greater volume of pattern 3, would be GG2 and would receive less aggressive treatment than a patient with a small amount of GP4 and no GP3 (GG4).

There is increasing evidence that among patients without pattern 5, the absolute length of GP4 on biopsy is a stronger predictor of oncologic outcomes than either the overall Gleason Score or the percentage of GP4 [3]. Recent data also indicate that once the burden of GP4 is accounted for, the presence of GP3 does not improve discrimination [4]. This suggests that the length of GP4 (in millimeters) on biopsy specimens should be the key determinant for risk stratification and treatment selection in men with GG2–4 disease.

However, such an approach cannot be implemented at the current time because there is important variation between pathologists in GP4 quantification. Andolfi et al. [5] reported a four-fold difference in median GP4 between pathologists at two high-volume academic centers seeing extremely similar patients. Bernhardt et al. examined the reasons for such variation by surveying 304 pathologists, identifying 18 distinct approaches to measuring tumor length and GP4 extent, with considerable variability in the resulting tumor measurements [6].

To our knowledge, there have been no attempts to systematically compare ways to quantify GP4 to determine an optimal approach, which we define as that which most accurately predicts oncologic outcomes. This may be because it is infeasible to ask pathologists to measure GP4 on a suitably large cohort of biopsy specimens and then repeat those measurements numerous times using different approaches.

Recent advances in machine learning (ML) have demonstrated computerized detection and grading of prostate cancer from digitalized biopsy slides [7]. This raises the possibility that ML could quantify cancer, and therefore, GP4 extent using multiple different approaches enabling direct comparison of their prognostic performance. In this study, we explore proof-of-principlefor such an approach. If successful, we would repeat our study on a larger, multi-institutional cohort to determine an optimal method for quantifying GP4.

## Methods

We identified 7,084 patients who had systematic prostate biopsy and underwent RP at MSKCC from 2012 to 2024. Although some patients had biopsies performed in-house, the majority underwent biopsy at outside institutions, with histologic slides submitted to MSKCCfor pathologic review. As GPU requirements increase with the number of cores examined, we selected a cohort of convenience from patients with a range of 2 to 6 positive cores.1,046 patients who had evidence of GP4 by ML constituted the cohort of the study. Cores from targeted biopsies were not included because it is unclear how to combine GP4 lengths from multiple cores in the same lesion.

We defined 15 approaches to GP quantification. We first distinguish between measuring all foci of cancer and the (disfavored) approach of measuring only the largest focus. When measuring all foci, we then used different approaches to incorporating the gaps between foci, that is, the interfocal stroma. These were either ignored or included depending on one of five inclusion criteria, namely if the interfocal stroma was: less than 3 mm; less than 1 mm; smaller than the combined lengths of the two adjacent foci; smaller than twice the combined lengths of the two adjacent foci; smaller than the shorter of the two adjacent foci.

Digitized biopsy slides were analyzed using software developed by the existing PAIGE-AI software to identify regions of GP3 and GP4. A new PAIGE-AI ML algorithm was developed and used to measure the lengths of individual pattern 4 foci directly from the digital images. This linear length of GP4 is a calculated value, not routinely stated in pathology reports. In our analysis, we assessed GG as determined by the proportions of Gleason patterns 3 and 4 identified by the ML, as well as GG assigned by the pathologist. For cores with GG2 and GG3, pathologists measured the cancer in mm and provided a percentage GP4. Length of GP4 for the core was obtained by multiplying those two measures. For cores with GG4, the entire length of cancer provided is presumed to be GP4.

We defined total GP4 length as the sum of GP4 lengths across all cancer-bearing cores with GG2-4. Measurements were repeated using the percentage of GP4 within each focus. Finally, we included a pixel-based area metric that ignored lengths and interfocal stroma, thus is not utilized in routine practice. Details of the various quantification methods, along with illustrative examples, are shown in the Supplementary Appendix.

To determine which measurement strategy had the greatest discrimination, area-under-the-curve (AUC) was estimated for each measurement combination for the primary outcome of adverse pathology, defined as seminal vesicle invasion (SVI) or nodal involvement (LNI). Pairwise comparisons of quantification approaches were performed using DeLong’s test [8]. We hypothesized that the rank order of different GP4 quantification approaches would be invariant to endpoint; it would not be the case that, for instance, incorporation of interfocal stroma is necessary to predict adverse pathology but not biochemical recurrence (BCR). To test this hypothesis, we evaluated the same comparisons for advanced disease including extraprostatic extension (EPE) and separately, for BCR. For BCR, we fitted Cox proportional hazards models and quantified discrimination using the concordance index, with 95%CI calculated by bootstrap.

We hypothesized that, if ML was a good tool to explore GP4 measurement, we should be able to replicate some other results reported in the literature. These include the discrimination of total length GP4 being superior to Gleason Score and %GP4; pattern 3 being non-predictive once pattern 4 is known; and clinical predictors (PSA, clinical stage, and total number of positive cores) adding little additional predictive value to the total length GP4. These hypotheses were tested using multivariable logistic regression models and discrimination of these multivariable models were calculated and corrected for optimism using 10-fold cross-validation with 1,000 iterations.

All analyses were conducted using R version 4.5.0 with the tidyverse (v2.0.0) and gtsummary (v2.2.0) packages [9].

## Results

We selected 1,046 patients from August 2014 to March 2023 with GG2-4 on systematic biopsy who underwent RP. Patients had either systematic biopsy or both systematic and targeted biopsy with no GP4 detected in the targeted cores. There were 293 patients excluded because they had more than one core per slide and 25 excluded who had pattern 5 defined by the PAIGE-AI algorithm. Of the remaining 753, analysis of 27 took excessive GPU time leaving a final analytic cohort of 726 patients. Overall, Table 1 shows that patients were relatively equally distributed across categories defined by the total number of positive biopsy cores.

**Table 1.** Patient and disease characteristics. Values are expressed as median (IQR) for continuous variables and frequency (%) for categorical variables.

| Characteristic | N = 726 |
| --- | --- |
| Age | 62 (57, 68) |
| Clinical Stage |  |
| ≤T1 | 465 (69%) |
| T2A | 125 (19%) |
| T2B | 51 (7.6%) |
| T2C | 5 (0.7%) |
| ≥T3 | 29 (4.3%) |
| Unknown | 51 |
| Number of Positive Biopsy Cores |  |
| 2 | 141 (19%) |
| 3 | 162 (22%) |
| 4 | 169 (23%) |
| 5 | 147 (20%) |
| 6 | 107 (15%) |
| Total PSA (ng/mL) | 6.3 (4.8, 8.7) |
| Unknown | 7 |
| Highest Biopsy Grade group assessed by ML |  |
| 2 | 486 (67%) |
| 3 | 183 (25%) |
| 4 | 57 (7.9%) |
| Adverse Pathology at RP (SVI or LNI) | 100 (14%) |
| Seminal Vesical Invasion | 51 (7%) |
| Lymph Node Invasion | 73 (10%) |
| Extraprostatic Extension | 324 (45%) |

Average GP4 lengths for each quantification metric, separately by GG, are shown in Table 2 and Supplementary Table 1. GP4 increases with GG but is relatively similar between quantification methods other than for the “largest focus only” method where, as expected, lengths are shorter. The different methods for incorporating interfocal stroma are highly correlated (>0.99). This appears to be because in most slides, there is either only one focus, or interfocal stroma is either very large (and thus excluded by all tested methods) or small (therefore included by all methods).

**Table 2.**
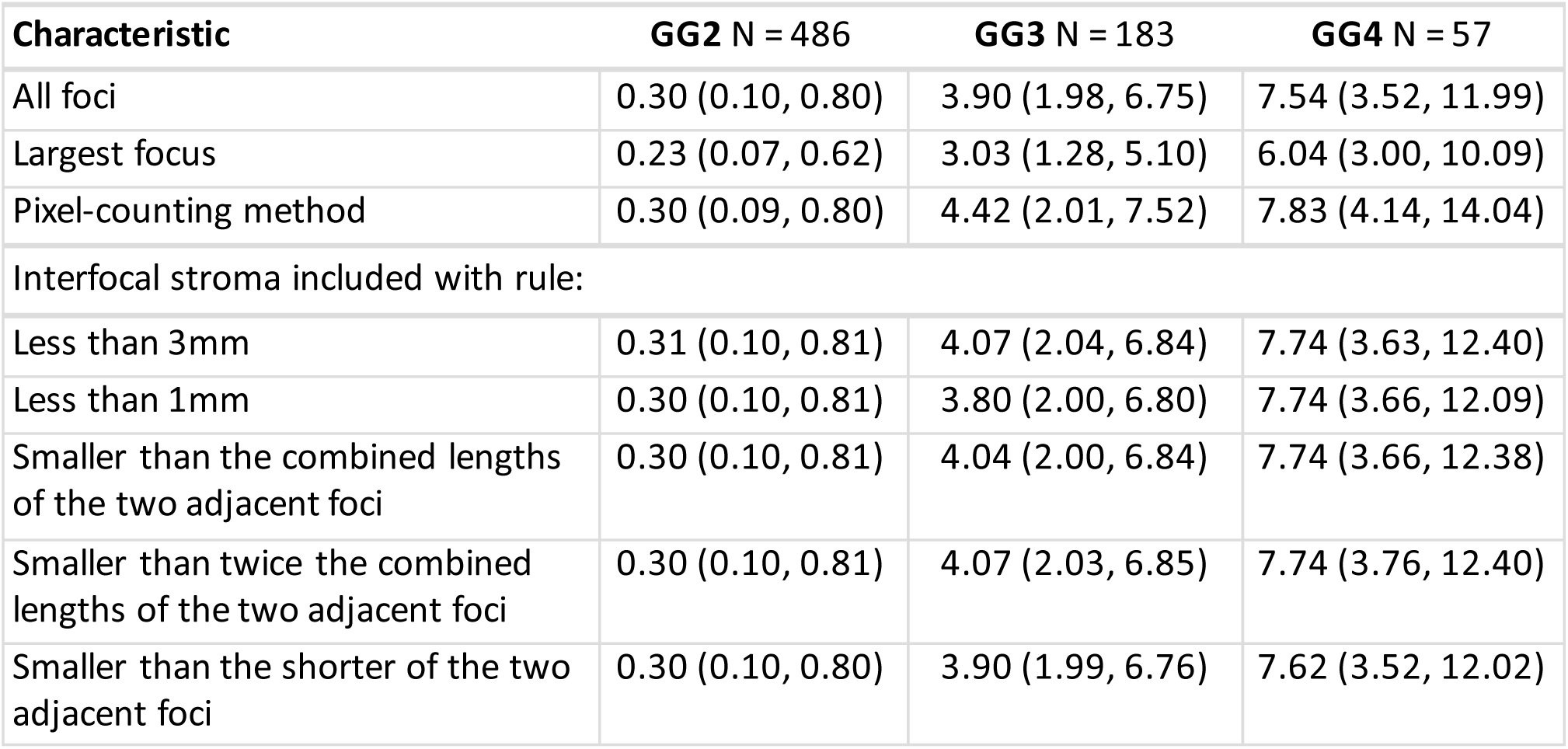
Gleason pattern 4 lengths (mm) measured by each of the seven interfocal stroma measurements by biopsy Grade Group, using tissue as the denominator (data for where cancer is the denominator shown in Supplementary Table 1). Values are expressed as median (IQR).

| Characteristic | GG2 N = 486 | GG3 N = 183 | GG4 N = 57 |
| --- | --- | --- | --- |
| All foci | 0.30 (0.10, 0.80) | 3.90 (1.98, 6.75) | 7.54 (3.52, 11.99) |
| Largest focus | 0.23 (0.07, 0.62) | 3.03 (1.28, 5.10) | 6.04 (3.00, 10.09) |
| Pixel-counting method | 0.30 (0.09, 0.80) | 4.42 (2.01, 7.52) | 7.83 (4.14, 14.04) |
| Interfocal stroma included with rule: |  |  |  |
| Less than 3mm | 0.31 (0.10, 0.81) | 4.07 (2.04, 6.84) | 7.74 (3.63, 12.40) |
| Less than 1mm | 0.30 (0.10, 0.81) | 3.80 (2.00, 6.80) | 7.74 (3.66, 12.09) |
| Smaller than the combined lengths of the two adjacent foci | 0.30 (0.10, 0.81) | 4.04 (2.00, 6.84) | 7.74 (3.66, 12.38) |
| Smaller than twice the combined lengths of the two adjacent foci | 0.30 (0.10, 0.81) | 4.07 (2.03, 6.85) | 7.74 (3.76, 12.40) |
| Smaller than the shorter of the two adjacent foci | 0.30 (0.10, 0.80) | 3.90 (1.99, 6.76) | 7.62 (3.52, 12.02) |

Discrimination for advanced stage (SVI or LNI) is shown in Table 3. As expected for a proof-of-principle study with a limited sample size, 95%CI are wide, precluding definitive conclusions. Nonetheless, several findings are of note. First, inclusion of all foci gives better discrimination than the “largest focus only” method which is disfavored in guidelines [10] . Second, we did not see evidence that incorporating interfocal stroma improved discrimination. Third, the method with the highest discrimination was the pixel-counting method which converts pixel area counts to length measurements. Total length GP4 was superior to GG (p=0.069 for the comparison with “all foci”; p=0.011 for the comparison with the pixel counting method). Results were virtually identical when advanced stage included EPE (Supplementary Table 2) and BCR (Table 4), supporting the hypothesis that rank ordering of GP4 quantification methods is independent of endpoint.

**Table 3.**
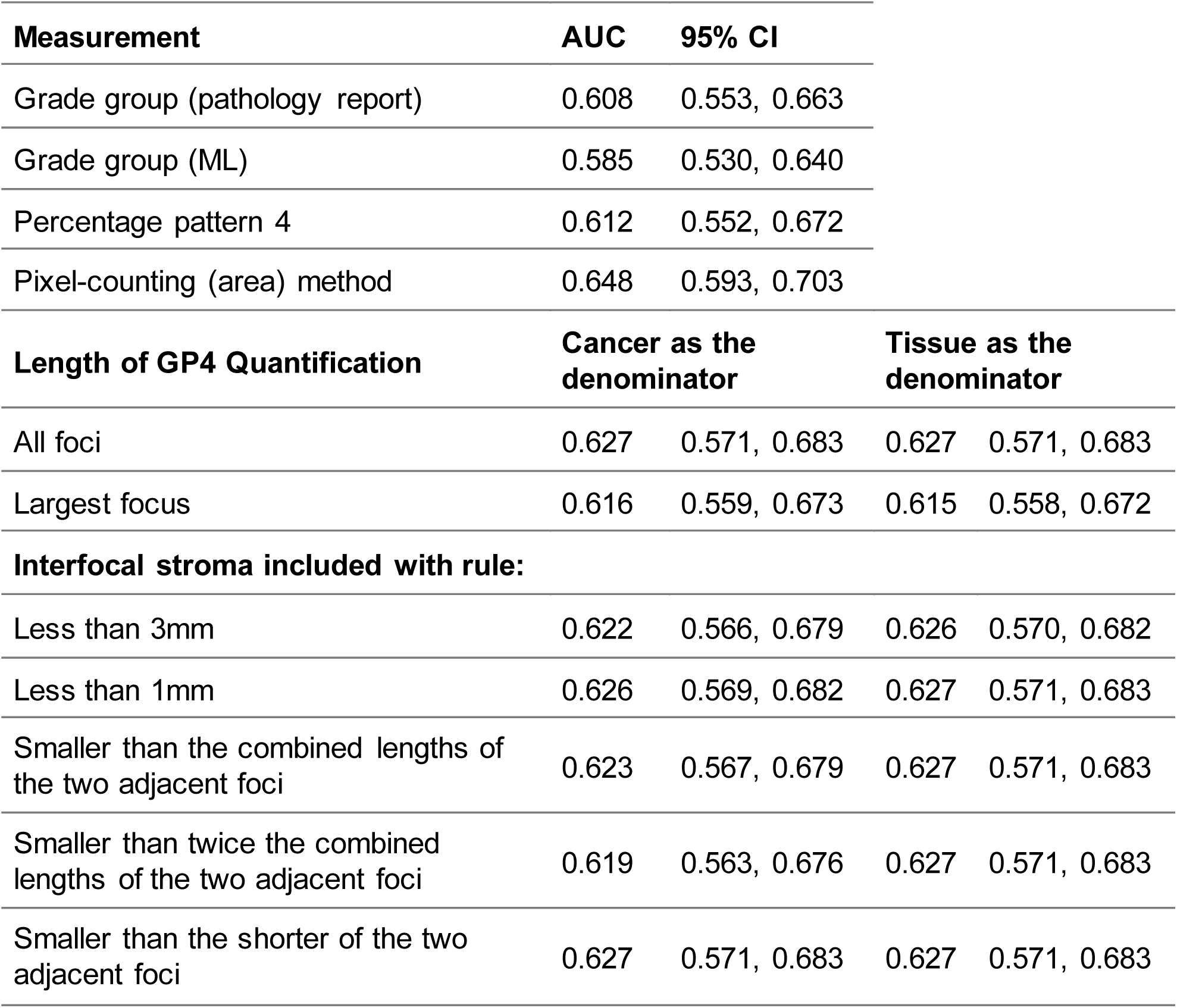
Discrimination (AUC) of advanced disease (SVI or LNI) for each of the 15 measurement types as well as Gleason score.

**Table 4.** Discrimination (C-Index) of BCR by each of the 15 measurement types.

| Measurement | C-index | 95% CI |  |  |
| --- | --- | --- | --- | --- |
| Grade group (pathology report) | 0.597 | 0.567, 0.668 |  |  |
| Grade group (ML) | 0.593 | 0.566, 0.681 |  |  |
| Percentage pattern 4 | 0.602 | 0.571, 0.687 |  |  |
| Pixel counting (area) method | 0.620 | 0.577, 0.660 |  |  |
| Length of GP4 Quantification | Cancer as the denominator |  | Tissue as the denominator |  |
| All foci | 0.610 | 0.566, 0.649 | 0.609 | 0.565, 0.649 |
| Largest focus | 0.603 | 0.560, 0.644 | 0.604 | 0.560, 0.644 |
| Interfocal stroma included with rule: |  |  |  |  |
| Less than 3mm | 0.611 | 0.567, 0.651 | 0.609 | 0.565, 0.649 |
| Less than 1mm | 0.610 | 0.565, 0.649 | 0.609 | 0.565, 0.649 |
| Smaller than the combined lengths of the two adjacent foci | 0.610 | 0.565, 0.649 | 0.609 | 0.565, 0.649 |
| Smaller than twice the combined lengths of the two adjacent foci | 0.610 | 0.566, 0.648 | 0.609 | 0.566, 0.650 |
| Smaller than the shorter of the two adjacent foci | 0.610 | 0.566, 0.650 | 0.609 | 0.565, 0.649 |

Discrimination for the pathologist-assigned GG was slightly better than for ML-assigned GG (0.608 vs. 0.585) confirming that while ML is adequate for research purposes, it is less accurate than specialist uropathologists. The discrimination for pathologist Gleason Score is lower than reported in other data sets, likely because choosing a convenience sample of patients with a lower number of cores leads to a more homogenous sample.

We replicated prior findings that, once the amount of GP4 is known, GP3 is non-predictive (p=0.1 for SVI/LNI). Among clinical predictors, the total number of positive cores was likewise non - predictive for SVI/LNI (p>0.9). We did find PSA (p<0.001) and clinical stage (p<0.001) were associated with SVI/LNI, but adding clinical predictors did not improve discrimination as cross-validated AUC was 0.636 (95% CI 0.570, 0.703) vs. 0.635 (95% CI 0.576, 0.695) with GP4 alone.

Findings were consistent for BCR, as GP3 remained non-predictive once GP4 was accounted for. Cross-validated discrimination for BCR was unchanged by adding clinical predictors.

## Discussion

Our findings indicate that ML can be used to quantify cancer and consequently GP4 on digitized biopsy slides using different approaches. This allowed us to efficiently process a large number of slides and directly compare different GP4 metrics to identify those with the strongest discriminative performance for predicting oncologic outcomes.

Proof-of-principle would be demonstrated if our results replicated prior findings, specifically: total length of GP4 in mm is a stronger predictor than the GG; once the extent of GP4 is known, the amount of GP3 does not improve prediction; clinical predictors contribute less prognostic information than GP4 burden. Proof-of-principle would also be demonstrated if measuring all tumor foci yielded better discrimination than the disfavored approach of measuring only the largest focus.

Firstly, our finding that the total length of GP4 is a stronger predictor of oncologic outcomes than the GG is consistent with results of Scuderi et al. [11] who reported that the total length of pattern 4 offered superior discrimination for EPE, SVI, and LNI compared with %GP4 or GG in a cohort of 1,171 men with GG2-4 prostate cancer undergoing RP. Pickersgill et al. [12] similarly reported higher discrimination for total GP4 length compared with GG in predicting adverse pathology (AUC 0.779 vs 0.658; p < 0.001) among 2,499 patients with GG2-4 disease. Olivier et al. [13] found in a cohort of 446 GG2-4 surgically treated patients with long term follow-up that the overall volume of GP4 on biopsy specimens offered superior discrimination to GG or %GP4 for predicting distant metastasis or BCR.

Secondly, we were able to replicate findings that GP3 does not improve discrimination once quantification of GP4 is known. Vickers et al. [4] found in multivariable model among patients with GG2 prostate cancer on biopsy, the addition of GP3 extent to the total length of GP4 was not associated with adverse pathology (EPE, SVI, LNI). Likewise, Pickersgill et al. [12], reported that the addition of GP3 length did not improve discrimination compared with total GP4 length alone for predicting adverse pathology (AUC 0.770 for combined pattern 4 and pattern 3 vs 0.783 for mm pattern 4 alone). Olivier et al. [13] observed that incorporating the extent of GP3 to the total length of GP4 on prostate biopsy fails to improve discrimination for BCR (c-index 0.706 vs 0.703) or metastatic disease (c-index 0.771 vs 0.772).

Thirdly, we found that although PSA (p < 0.001) and clinical stage (p < 0.001) were independently associated with clinical outcomes in multivariable analysis, incorporating these clinical predictors alongside total length GP4 failed to improve discrimination. The combined model (c-index 0.620) did not perform better than GP4 alone (c-index 0.618). Pickersgill et al. [12] reported that adding clinical predictors to the amount of GP4 on biopsy reduced discrimination for adverse pathology (AUC 0.784 vs 0.776) and BCR (c-index 0.707 vs 0.677).

Lastly, the “largest focus” method is rarely used by pathologists, as measuring only the longest focus in a core containing multiple discontinuous foci likely underestimates true tumor extent [6]. As a negative example in our analysis, it yielded the lowest discriminative performance (c-index 0.615) compared with methods that accounted for all tumor foci as expected.

There is a dearth of papers examining the optimal method for quantifying GP4, and the few that exist are limited by small cohort sizes. For example, Karam et al. [14] reported that including interfocal stroma when measuring discontinuous tumor foci increased the correlation with radical prostatectomy findings in a cohort of 109 patients. Similarly, Arias-Stella et al. [15] evaluated a cohort of 40 patients with discontinuous tumor foci on prostate biopsy and correlated these findings with RP specimens. They reported that 78% of cases represented a single irregularly shaped tumor focus, suggesting interfocal stroma should not be excluded, as discontinuous foci more likely reflect a single tumor sampled at multiple points by the biopsy needle rather than truly distinct lesions. It remains unclear whether these studies were conducted with prespecified hypotheses and statistical methods regarding the inclusion of interfocal stroma.

Notably, at least in part based on these findings, current guidelines recommend the inclusion of interfocal stroma. For instance, the College of American Pathologists advises measuring a tumor focus from one end to the other, including intervening benign tissue [10]. In contrast, our analysis found no evidence that incorporating interfocal stroma improves discrimination. Although, the wide confidence intervals in our study preclude definitive conclusions, the discordance between our results and guideline recommendations highlights the need for further investigation to clarify the role of interfocal stroma in GP4 quantification. Such investigation would require large sample sizes and clearly prespecified hypotheses and analyses.

Several methodological challenges had to be overcome before we were able to move from ML recognition of Gleason patterns to measurement of GP4. A notable limitation was the inability of the ML algorithm to distinguish between biopsy cores and levels on a single slide. During histopathologic processing, biopsy fragments are sectioned at multiple levels to allow evaluation of the same core across different depths. This process may result in core fragmentation, producing multiple tissue objects on a single slide. The ML was unable to determine whether these objects represented different levels of the same core or distinct fragments of a single discontinuous core. To address this limitation, an investigator reviewed each slide to confirm the correct number of cores and levels. This process was less time-consuming than a full pathological review, requiring only a brief visual inspection of the tissue fragments on each slide. Other technical challenges encountered during the development of the ML, and the corresponding solutions are described in the Supplementary Appendix.

A limitation of our study is the inclusion of cribriform glands. The PAIGE-AI algorithm classifies cribriform glands as GP4, as they represent one of the architectural patterns encompassed within pattern 4 according to current ISUP grading criteria. However, invasive cribriform architecture has been recognized as an independent prognostic factor for oncologic outcomes in prostate cancer [16]. Thus, evaluation of cribriform architecture would be considered in future studies.

In conclusion, our preliminary data demonstrates the feasibility of using ML as a research tool to quantify GP4 using multiple different measurement approaches. As such, we plan to repeat this study on a much larger and multinstitutional study to improve the precision of our findings and make firm recommendations for pathologic practice.

## Data Availability

All data produced in the present study are available upon reasonable request to the authors

---

**Supplementary Table 1.**
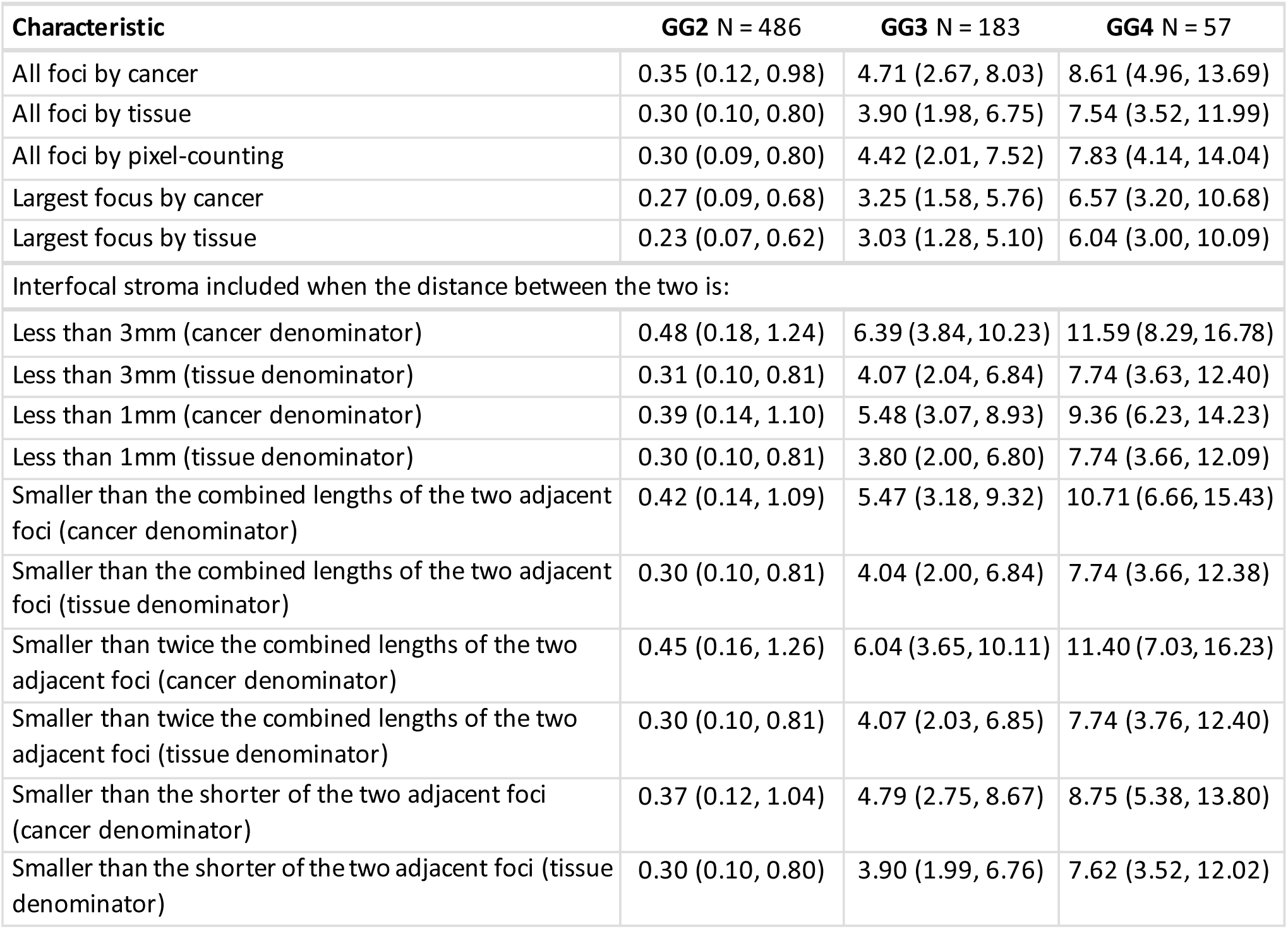
Pattern 4 lengths (mm) by proportion of either cancer or tissue for each of the 15 measurement types by biopsy Gleason Grade. Values are expressed as median (IQR).

| Characteristic | GG2 N = 486 | GG3 N = 183 | GG4 N = 57 |
| --- | --- | --- | --- |
| All foci by cancer | 0.35 (0.12, 0.98) | 4.71 (2.67, 8.03) | 8.61 (4.96, 13.69) |
| All foci by tissue | 0.30 (0.10, 0.80) | 3.90 (1.98, 6.75) | 7.54 (3.52, 11.99) |
| All foci by pixel-counting | 0.30 (0.09, 0.80) | 4.42 (2.01, 7.52) | 7.83 (4.14, 14.04) |
| Largest focus by cancer | 0.27 (0.09, 0.68) | 3.25 (1.58, 5.76) | 6.57 (3.20, 10.68) |
| Largest focus by tissue | 0.23 (0.07, 0.62) | 3.03 (1.28, 5.10) | 6.04 (3.00, 10.09) |
| Interfocal stroma included when the distance between the two is: |  |  |  |
| Less than 3mm (cancer denominator) | 0.48 (0.18, 1.24) | 6.39 (3.84, 10.23) | 11.59 (8.29, 16.78) |
| Less than 3mm (tissue denominator) | 0.31 (0.10, 0.81) | 4.07 (2.04, 6.84) | 7.74 (3.63, 12.40) |
| Less than 1mm (cancer denominator) | 0.39 (0.14, 1.10) | 5.48 (3.07, 8.93) | 9.36 (6.23, 14.23) |
| Less than 1mm (tissue denominator) | 0.30 (0.10, 0.81) | 3.80 (2.00, 6.80) | 7.74 (3.66, 12.09) |
| Smaller than the combined lengths of the two adjacent foci (cancer denominator) | 0.42 (0.14, 1.09) | 5.47 (3.18, 9.32) | 10.71 (6.66, 15.43) |
| Smaller than the combined lengths of the two adjacent foci (tissue denominator) | 0.30 (0.10, 0.81) | 4.04 (2.00, 6.84) | 7.74 (3.66, 12.38) |
| Smaller than twice the combined lengths of the two adjacent foci (cancer denominator) | 0.45 (0.16, 1.26) | 6.04 (3.65, 10.11) | 11.40 (7.03, 16.23) |
| Smaller than twice the combined lengths of the two adjacent foci (tissue denominator) | 0.30 (0.10, 0.81) | 4.07 (2.03, 6.85) | 7.74 (3.76, 12.40) |
| Smaller than the shorter of the two adjacent foci (cancer denominator) | 0.37 (0.12, 1.04) | 4.79 (2.75, 8.67) | 8.75 (5.38, 13.80) |
| Smaller than the shorter of the two adjacent foci (tissue denominator) | 0.30 (0.10, 0.80) | 3.90 (1.99, 6.76) | 7.62 (3.52, 12.02) |

**Supplementary Table 2.** Discrimination (AUC) of the alternative definition of advanced disease where EPE is also considered advanced disease by each of the 15 measurement types.

| Measurement | AUC | 95% CI |  |  |
| --- | --- | --- | --- | --- |
| Grade group (pathology report) | 0.577 | 0.540, 0.614 |  |  |
| Grade group (ML) | 0.536 | 0.502, 0.571 |  |  |
| Percentage pattern 4 | 0.559 | 0.517, 0.601 |  |  |
| Pixel counting (area) method | 0.612 | 0.571, 0.653 |  |  |
|  | Cancer as the denominator |  | Tissue as the denominator |  |
| All foci | 0.592 | 0.550, 0.633 | 0.596 | 0.555, 0.637 |
| Largest focus | 0.589 | 0.547, 0.630 | 0.591 | 0.549, 0.632 |
| **Interfocal stroma included with rule:** |  |  |  |  |
| Less than 3mm | 0.588 | 0.547, 0.629 | 0.595 | 0.554, 0.636 |
| Less than 1mm | 0.590 | 0.548, 0.631 | 0.596 | 0.555, 0.637 |
| Smaller than the combined lengths of the two adjacent foci | 0.588 | 0.547, 0.630 | 0.595 | 0.554, 0.637 |
| Smaller than twice the combined lengths of the two adjacent foci | 0.585 | 0.543, 0.626 | 0.595 | 0.554, 0.636 |
| Smaller than the shorter of the two adjacent foci | 0.591 | 0.550, 0.632 | 0.596 | 0.555, 0.637 |

## Supplementary methods

Digitized prostate biopsy slides were analyzed using a machine-learning algorithm (PAIGE-AI) to quantify tissue area, cancer area, and Gleason pattern 4 (GP4). Size-thresholding was applied to exclude isolated regions smaller than 700 µm². Each biopsy core may be sectioned at multiple histologic levels; these levels appear on a slide as separate objects, all corresponding to the same physical biopsy core.

Object-level measurements, including core length and GP4 length, were generated for each level. To exclude tissue fragments and artifacts, object-level core lengths within each slide were reviewed; small objects that deviated substantially from the dominant core length were classified as fragments and removed. Slides with extensive fragmentation, artifacts, or no measurable GP4 were excluded.

For each biopsy core, GP4 length was calculated as the average GP4 length across its sectioned levels. Patient-level GP4 burden was then computed by summing the average GP4 length across all included biopsy cores for that patient.

To illustrate the fifteen different GP4 quantification approaches, we use a single biopsy core example below, where light blue is normal tissue, dark blue is pattern 3 and dark green is pattern 4. Three cancer foci are separated by interfocal stroma measuring 0.9 mm and 3 mm. The total cancer length is 7 mm, of which 5 mm is GP4. Each focus occupies approximately 50% of the core width.

We first distinguish between foci inclusion, where we either sum measurements across all foci or measure only the longest focus. In this case, the largest focus is 3 mm long, 2 mm of which is GP4 to give a 67% GP4 percentage. Using a cancer-based denominator, the GP4 fraction is 3 × 0.67, yielding 2.0 mm of GP4. The focus spans half the core such as that GP4 is 33% of the tissue, hence using a tissue as the denominator give 3 × 0.33 = 1.0 mm of GP4.

The next question is whether the interfocal stroma would be included under different inclusion rules. Interfocal stroma was included based on five rules: including interfocal stroma < 3 mm; 1 mm; shorter than the combined lengths of adjacent foci; shorter than twice the combined lengths; or shorter than the shorter adjacent focus.

Under these rules, the 0.9 mm interfocal stroma would be included by all five rules (<3 mm, <1 mm, shorter than the combined lengths of adjacent foci, shorter than twice the combined lengths, and shorter than the shorter adjacent focus). The 3 mm interfocal stroma would only be included by the two relative-length rules based on the combined lengths of the adjacent foci (shorter than the combined lengths and shorter than twice the combined lengths). Now that interfocal stroma is included under some rules, we next consider whether cancer or tissue is used as the denominator.

If cancer is used as the denominator, the calculation is as follows: the total cancer length is 7 mm. When the 0.9 mm interfocal stroma is included, the counted length becomes 7.9mm. The fraction of cancer that is GP4 is 5 ÷ 7. Multiplying the counted length by this fraction yields 5.6 mm of GP4. If both interfocal stromal segments are included, the counted length becomes 10.9 mm and multiplying by 5 ÷ 7 yields 7.8 mm of GP4.

If tissue is used as the denominator, the calculation changes. Ignoring interfocal stroma, the total tissue length is 7 mm. Of this tissue, 46.7% is GP4, calculated as (2mm × 0.5 + 1.5 mm × 1.0 + 1.5 mm × 0.5)/7 mm, yielding 3.25 mm of GP4. Including the 1 mm interfocal stroma increases the total tissue length to 8 mm, reducing the GP4 fraction to 40.6%, but the product remains 3.25 mm. Including both interfocal stromal segments increases the tissue length to 11 mm and reduces the GP4 fraction to 29.5%, again yielding mm of GP4.

In other words, when tissue is used as the denominator, explicitly calculating interfocal stroma does not change the GP4 estimate. Including interfocal stroma increases the counted length, but this is exactly offset by the corresponding decrease in the GP4 fraction, resulting in the same final value.

Finally, pixel areas were converted to microns using slide-specific microns-per-pixel (mpp) values, and the resulting GP4 area was converted to a linear extent by dividing by the average core width. This is equivalent to estimating the percentage of the core occupied by GP4 and multiplying by the total core length. This method considers variations in the width of a core over its length. If the width of the GP4 at each focus is the same as the average core width, the length calculated by this method will match the all-foci tissue-based approach. See Supplementary Figure 1 and 2.

## Methodologic challenges for ML measurement of GP4

1. The algorithm was designed to measure lengths by drawing a consistent end-to-end midline along each core. We noticed that in some cases, small tissue irregularities would cause the midline to deviate, leading to inaccurate measurements. Example shown in Supplementary Figure 3.
2. The algorithm quantified object lengths in pixels and converted them to microns using a predefined scale factor. However, we observed substantial discrepancies in the measured specimen lengths. Further investigation revealed that these inconsistencies were due to variations in slide magnification. To address this, we adjusted the pixel-to-micron conversion rate to account for magnification differences.
3. During the initial phases of the project, some fragments were recorded as having a total length of zero. Further investigation revealed that when a fragment of a biopsy core contained no GP4, the algorithm assigned a total length of zero to that entire object. This behavior resulted in an underestimation of the total tissue length for the core. For example, in Supplementary figure 4 we have 2 fragments of a biopsy core where the first fragment would have been considered a length of zero, thus underestimating the total tissue length by half.
4. We found large discrepancies in GP4 lengths across levels from the same core when interfocal stroma was included. After manual review, we observed that in several cases, the algorithm labeled scattered pixels of GP4 on a fragment, which distorted the measurements. To prevent these small detections from affecting the results, we implemented a minimum threshold for GP4 foci before they were included in the length calculations. Example shown in Supplementary Figure 5.
5. During histopathologic processing, biopsy fragments are sectioned at multiple levels to enable evaluation of the same core at varying depths. As a result, multiple representations of the same biopsy core may appear on a single slide. The machine learning model had difficulty distinguishing different levels of the same core from distinct cores present on the slide. For this, manual review was required to verify core identity and ensure accurate measurements. Example shown in Supplementary Figure 6.
6. In some cases, multiple biopsy fragments were present within a single biopsy core, which inflated the measured tissue or cancer size. Manual review was necessary to identify these instances and adjust the measurements by accounting for the number of fragments present. Example shown in Supplementary Figure 7.
7. Due to storage-related issues, the machine learning system occasionally saved biopsy slides containing multiple levels as separate images. As a result, these images were incorrectly treated as distinct biopsy fragments. Manual review was required to identify such cases and exclude duplicate measurements.
8. During histopathological processing, biopsy fragments may become fragmented, resulting in discontinuous foci. In such cases, manual review was required to determine whether fragmentation was so severe it would exclude the respective core from the analysis. Example shown in Supplementary Figure 8.

---

**Supplementary Figure 1.**
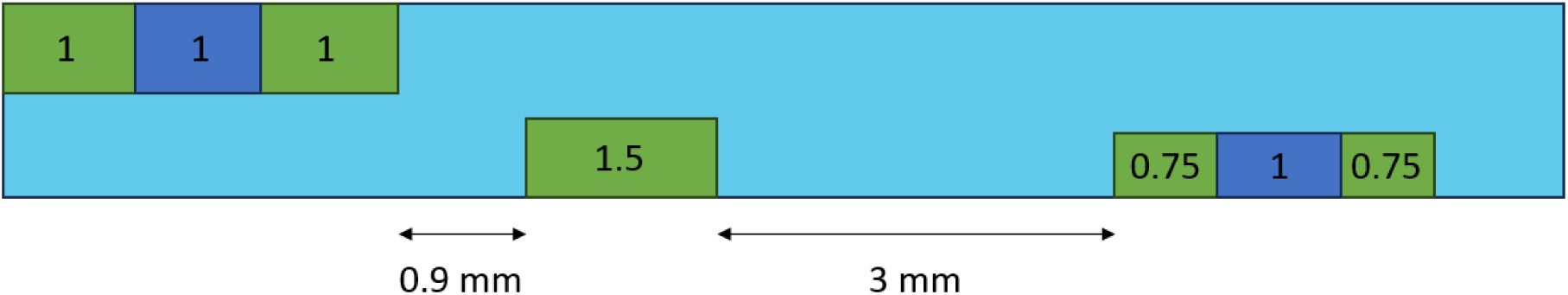
Scheme of a hypothetical biopsy core. Light blue: benign tissue. Dark Blue: Gleason pattern 3. Green: Gleason pattern 4. All values are given in mm.

**Supplementary Figure 2.**
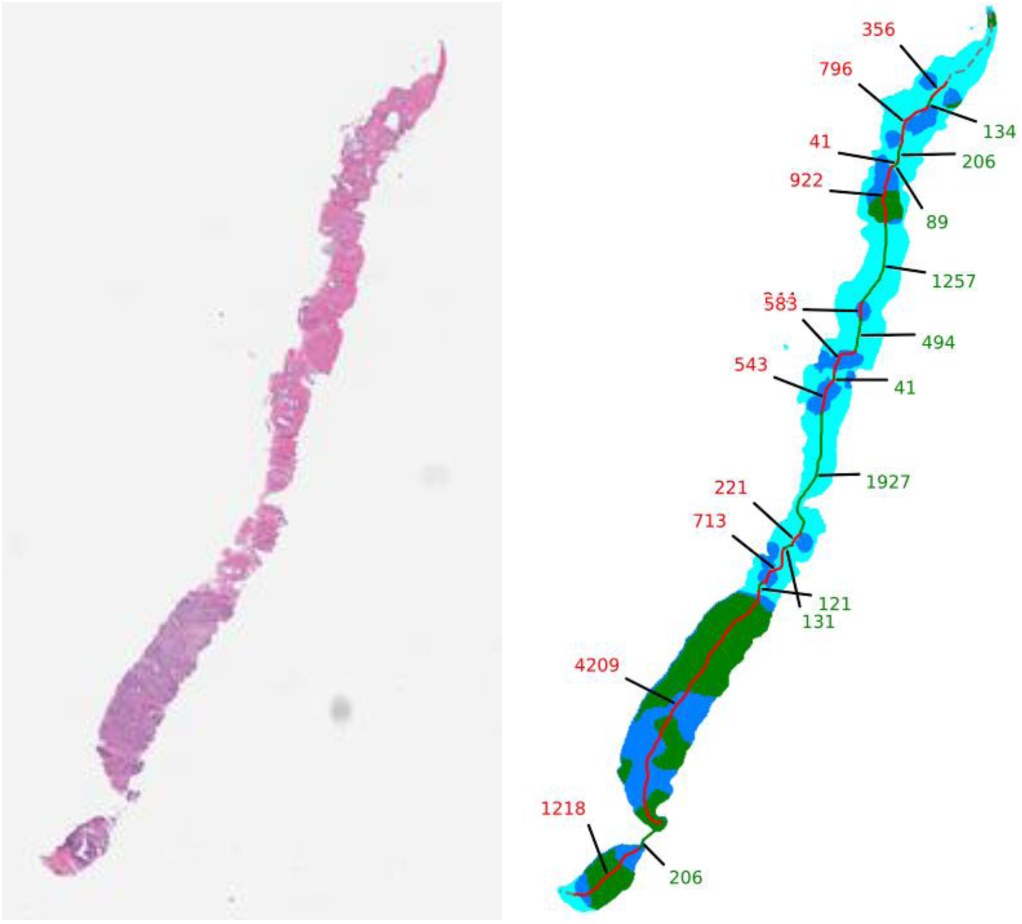
Left panel: Hematoxylin-eosin-stained biopsy slide from a patient with prostate cancer. Right panel: Digitized image of the corresponding biopsy slide. Light blue indicates benign tissue, dark blue indicates Gleason Pattern 3 (GP3), and dark green indicates Gleason Pattern 4 (GP4). The measured length (microns) of the cancer focus is highlighted and annotated in red and the measured length of interfocal stroma is annotated in green.

**Supplementary Figure 3.**
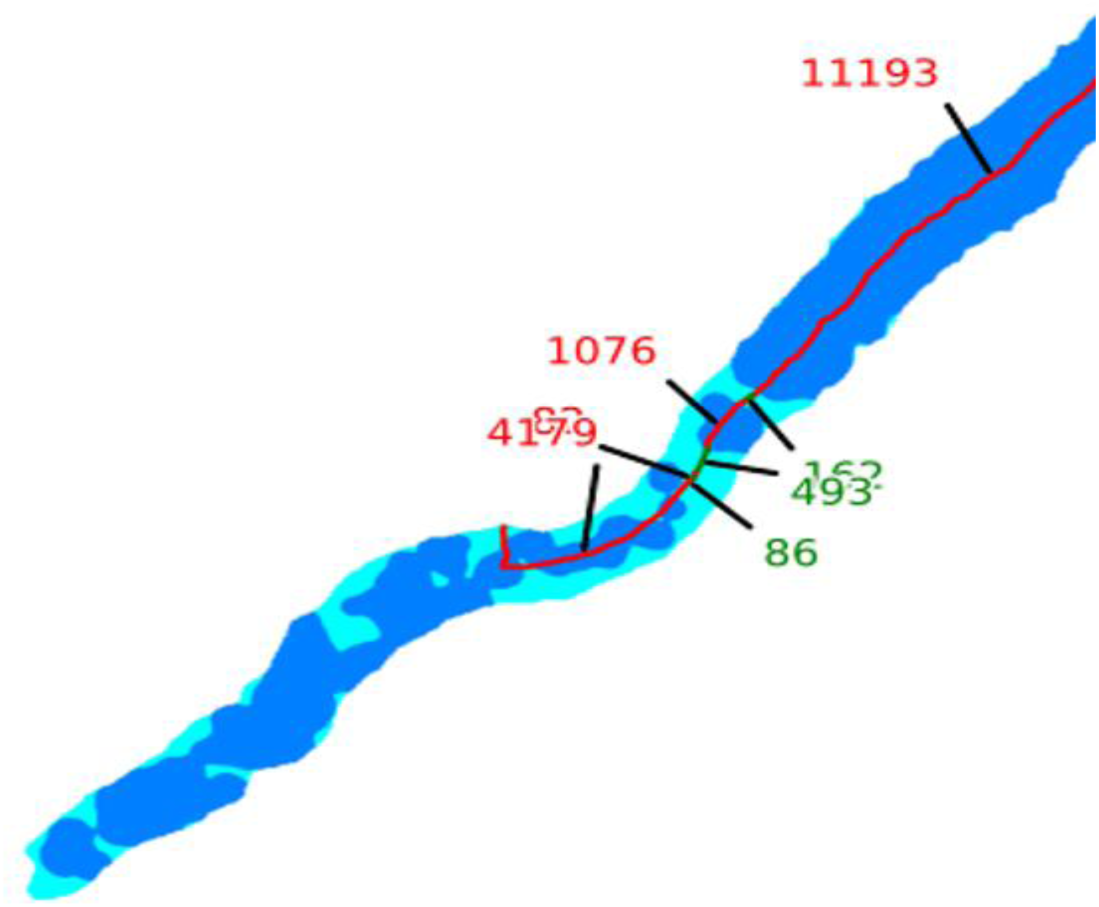
Example of biopsy slide with midline deviation.

**Supplementary Figure 4.**
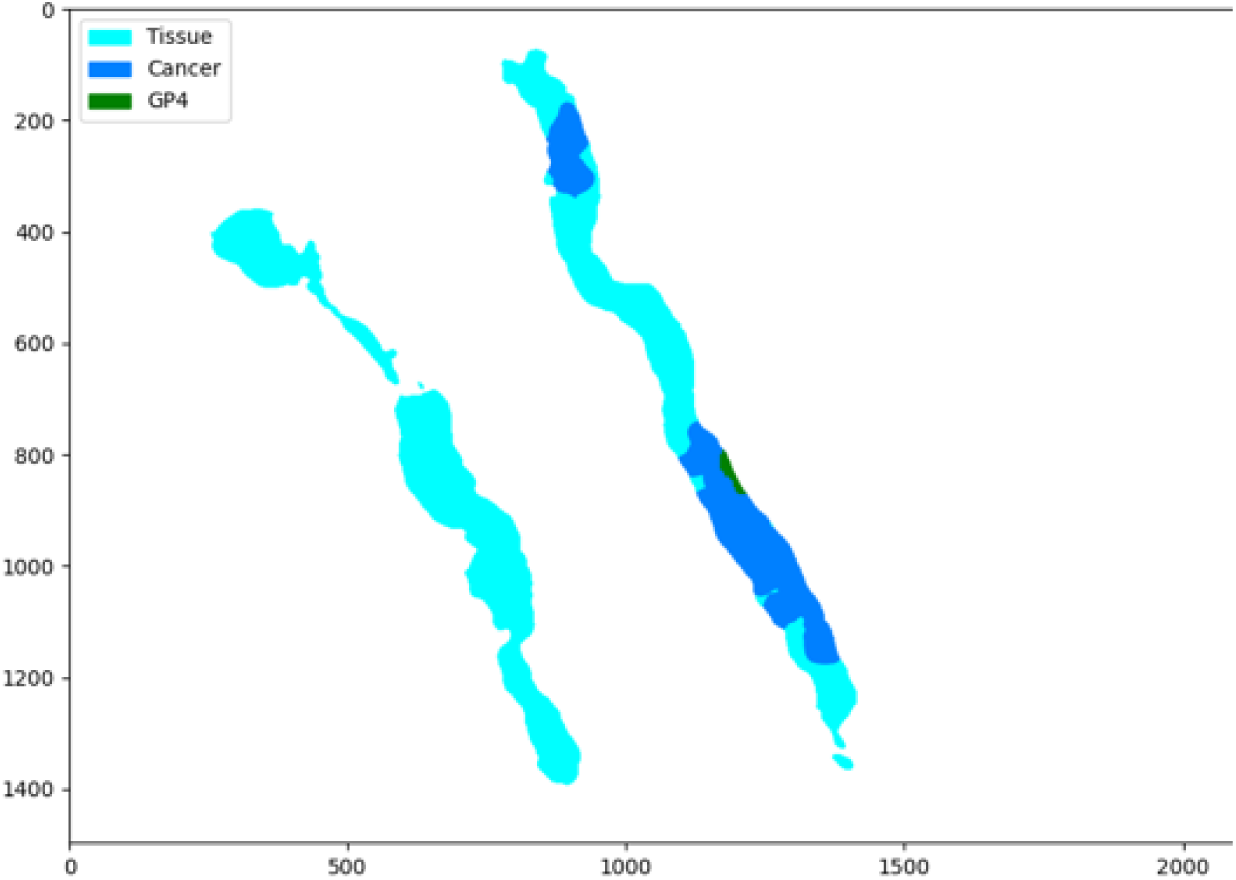
Example of biopsy slide with no pattern 4 on one core.

**Supplementary Figure 5.**
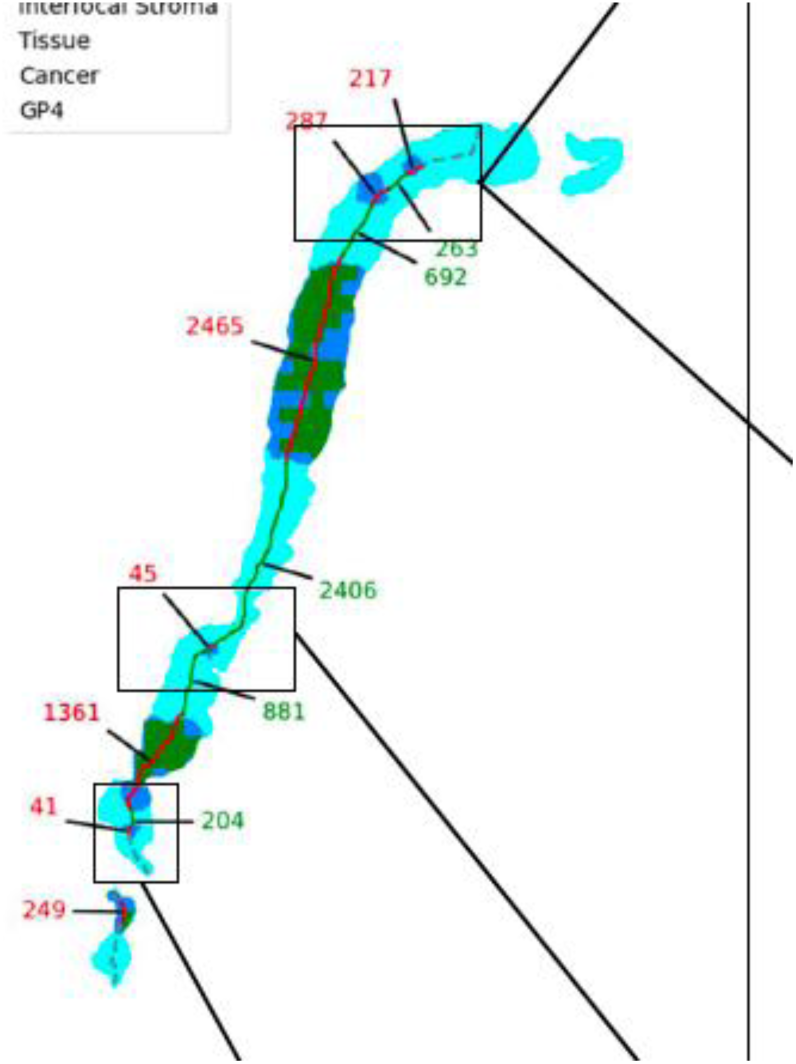
Example of biopsy slide with scattered pixels of pattern 4.

**Supplementary Figure 6.**
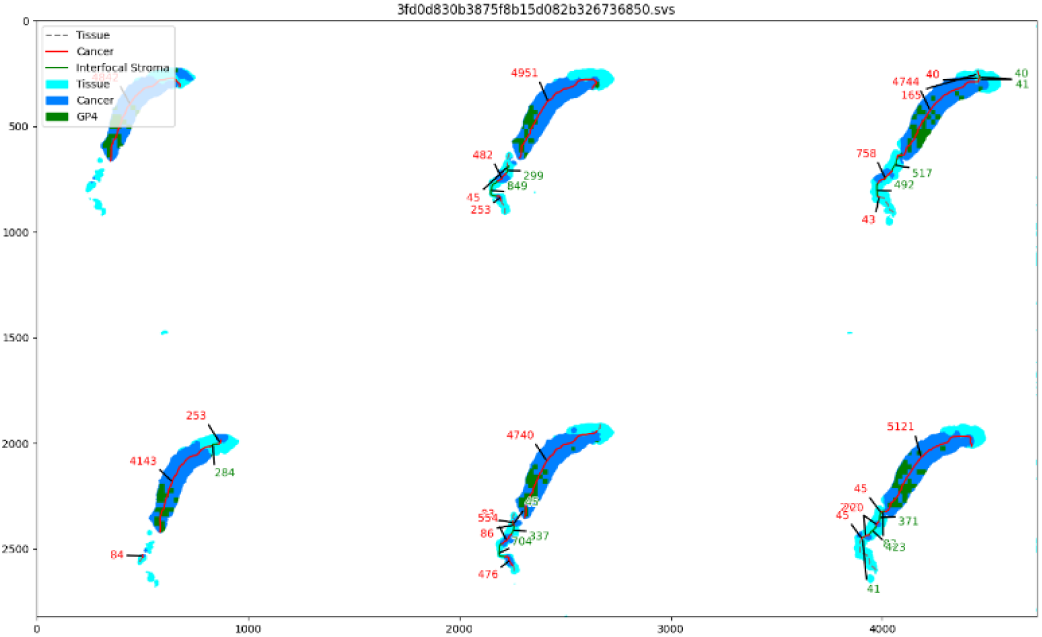
Example of biopsy slide with multiple different levels of the same core.

**Supplementary Figure 7.**
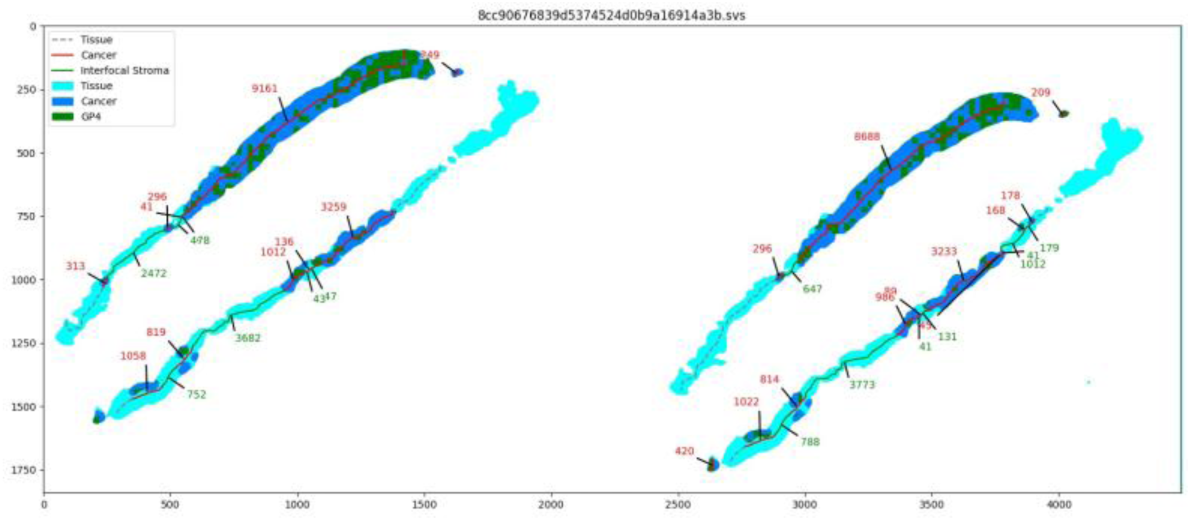
Example of biopsy slide with multiple cores and multiple levels

**Supplementary Figure 8.**
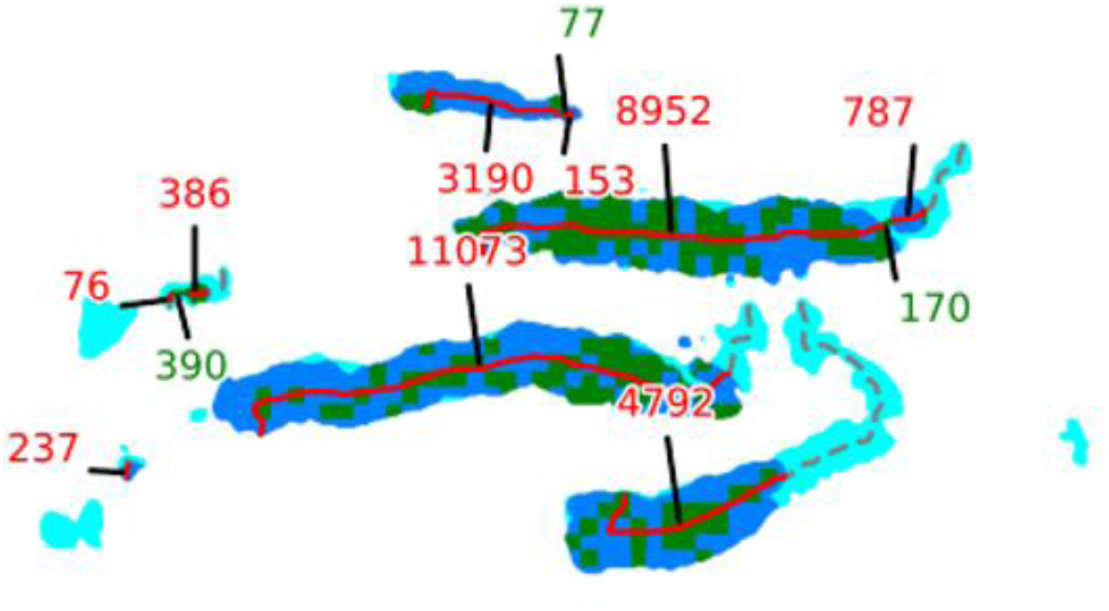
Example of a completely fragmented biopsy core that required exclusion from the analysis.

